# An emerging clade of Chikungunya West African genotype discovered in real-time during 2023 outbreak in Senegal

**DOI:** 10.1101/2023.11.14.23298527

**Authors:** Abdou Padane, Houriiyah Tegally, Yajna Ramphal, Ndiaye Seyni, Mariéma Sarr, Mame Matar Diop, Cyrille Kouligueul Diedhiou, Aminata Mboup, Ndèye Dieyna Diouf, Abdoulaye Souaré, Ndéye Diabou Diagne, Marilyne Aza-Gnandji, Ndèye Astou Dabo, Yacine Amet Dia, Ndeye Aminata Diaw, Nafissatou Leye, Papa Alassane Diaw, Ambroise Ahouidi, Badara Cissé, Abdoulaye Samba Diallo, Ousmane Diop, Abdou Aziz Diallo, Souadou Ndoye, Tomasz J. Sanko, Cheryl Baxter, Eduan Wilkinson, James E. San, Derek Tshabuila, Yeshnee Naidoo, Sureshnee Pillay, Richard Lessells, Khady Cissé, Abdoulaye Leye, Khalifa Ababacar Mbaye, Dramane Kania, Bachirou Tinto, Isidore Traoré, Sampawendé Thérèse Kagone, Abdoul Salam Ouedraogo, Robert J. Gifford, José Lourenço, Marta Giovanetti, Jennifer Giandhari, Tulio de Oliveira, Souleymane Mboup

## Abstract

Chikungunya (CHIKV) is a re-emerging endemic arbovirus in West Africa. Since July 2023, Senegal and Burkina Faso have been experiencing an ongoing outbreak, with over 300 confirmed cases detected so far in the regions of Kédougou and Tambacounda in Senegal, the largest recorded outbreak yet. CHIKV is typically maintained in a sylvatic cycle in Senegal but its evolution and factors contributing to re-emergence are so far unknown in West Africa, leaving a gap in understanding and responding to recurrent epidemics. We produced, in real-time, the first locally-generated and publicly available CHIKV whole genomes in West Africa, to characterize the genetic diversity of circulating strains, along with phylodynamic analysis to estimate time of emergence and population growth dynamics. A novel strain of the West African genotype, phylogenetically distinct from strains circulating in previous outbreaks, was identified. This suggests a likely new spillover from sylvatic cycles in rural Senegal and potential of seeding larger epidemics in urban settings in Senegal and elsewhere.

## Main Text

Chikungunya virus (CHIKV) is an arbovirus transmitted by mosquitoes of the genus *Aedes*. The disease it causes is associated with acute fever and musculoskeletal pain that can become chronic, with debilitating rheumatic disease in a substantial portion of infected individuals. The first detection of CHIKV in the world was in Tanzania in East Africa in 1952 ^1^. In Senegal, the first human case was identified in 1966 in Rufisque^2^. Since then, sporadic cases and epidemics have been detected in the country, where CHIKV circulates in sylvatic and urban cycles, with non-human primates acting as amplification hosts^3^. Spread among non-human primates is influenced by immune status, susceptible simian populations and the migration of infected sylvatic vectors into human-inhabited regions^3^. This potentially leads to inter-epizootic silence ranging between 3-5 years within the CHIKV transmission cycle. Recent recorded outbreaks in Senegal occurred in 2009/2010 and 2015^4^, often in the region of Kédougou, along with sporadic cases in 2021 and 2022, and an ongoing outbreak in 2023 (https://www.sante.gouv.sn/). Recorded CHIKV cases are likely underestimated due to limited diagnostic capacity in rural regions and overlapping clinical symptoms with other common febrile infections, including malaria.

CHIKV transmission dynamics and re-emergence patterns are not fully characterized in West Africa, leaving a severe gap in understanding and responding to recurrent outbreaks in the region. In Asia, Latin America, and other parts of Africa (Indian Ocean epidemic in 2005), CHIKV has previously caused large and often deadly epidemics^5–9^. It is therefore critical to fill gaps in the genomic characterization of CHIKV in West Africa, in real-time during outbreaks for public health response, and to understand the progression from initial emergence in rural communities to the potential of large outbreaks in urban settings. Additionally, CHIKV vaccines recently on the market are based on the East-Central-South-African (ECSA) genotype, which highlights the importance of contemporaneous genomic surveillance in regions where other lineages are known to circulate^10^.

In Senegal, in July 2023, a new case of CHIKV was notified in the region of Kédougou, particularly in Saraya. A month later, on 16 August 2023, cases had risen to 66, spreading to Kédougou city, Bandafassi and Ndiormi. By the end of October, there were 303 total confirmed CHIKV in Senegal; 57.8% of which were in the Kédougou region (Figure 1A). In the Kédougou health district, the most affected communities were Bandafassi, which accounted for over 50% of cases, particularly in the village of Syllacounda Bandafassi, followed by Tomborokoto (Bantaco) and Kédougou (Ndiormy and Dalaba). Subsequently, mass communication and localized vector control measures were implemented to curb the spread of the epidemic. The region also recorded 7 cases of CHIKV in 2021-2022 and is experiencing simultaneous circulation of dengue in 2023.

**Figure 1:**
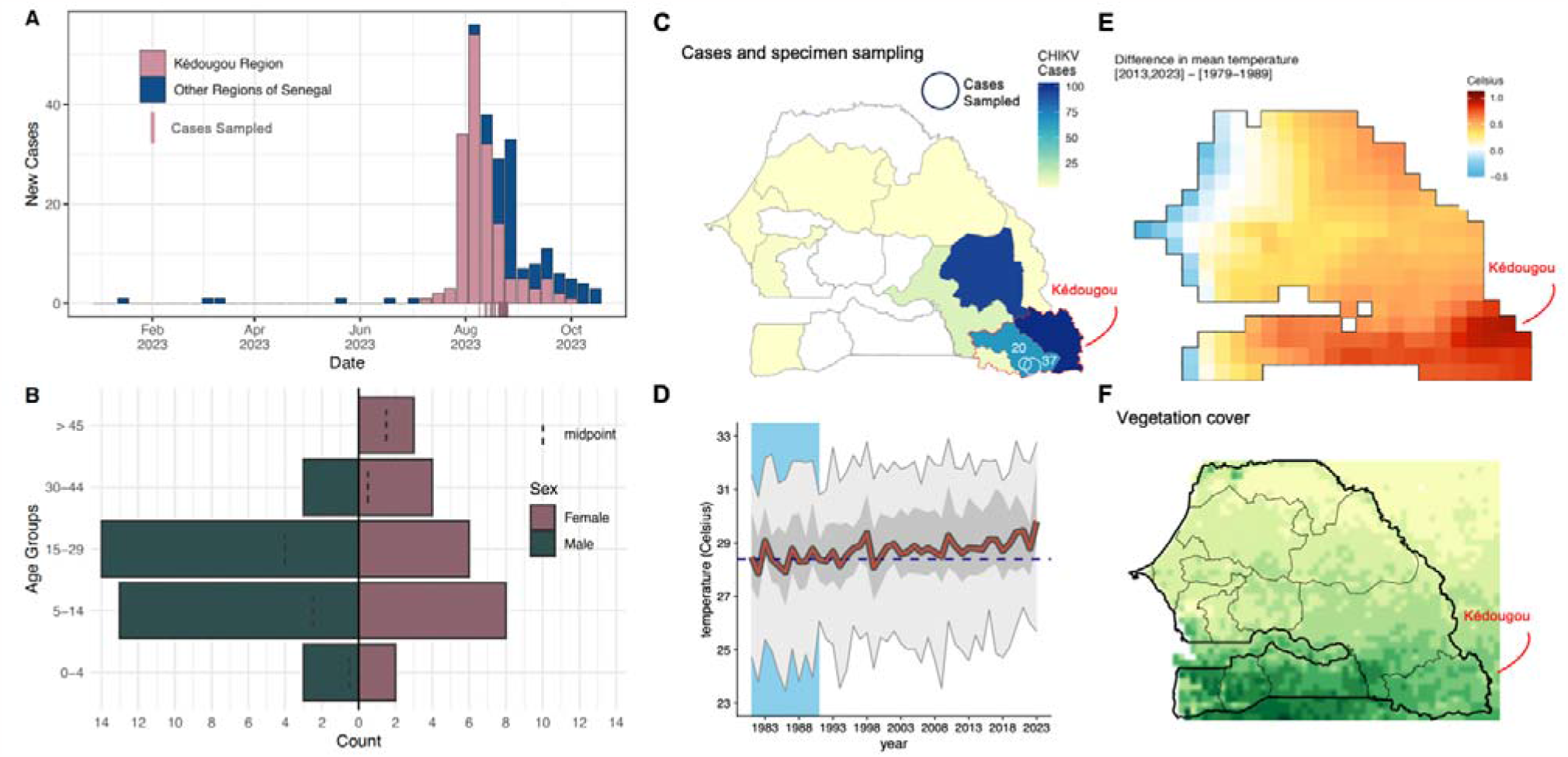
Spatiotemporal distribution of Chikungunya epidemiology and ecology in Senegal. A) Epidemiological curve of cases in Senegal and Kédougou region in 2023 and time-frame of case sampling. B) Demographic distribution of sampled cases. C) Map of CHIKV cases in Senegal in 2023, with the Kédougou region outlined in red. Sampling was performed in the Kédougou district, from Ndiormi (n=37) and Bandafassi (n=20). D) Increasing temperature trend in Senegal; the blue dashed line is the mean of the first 10 years of data (1979-1989, blue area), the red line is the annual mean, dark gray is the 50% percentile, and the light gray area are the minimum and maximum temperatures. E) Difference in mean temperature between the reference period of the first panel and the mean of the past 10 years (2013-2023). F) Vegetation cover (NDVI) in Senegal. Data is from 2018.

**Figure 2.**
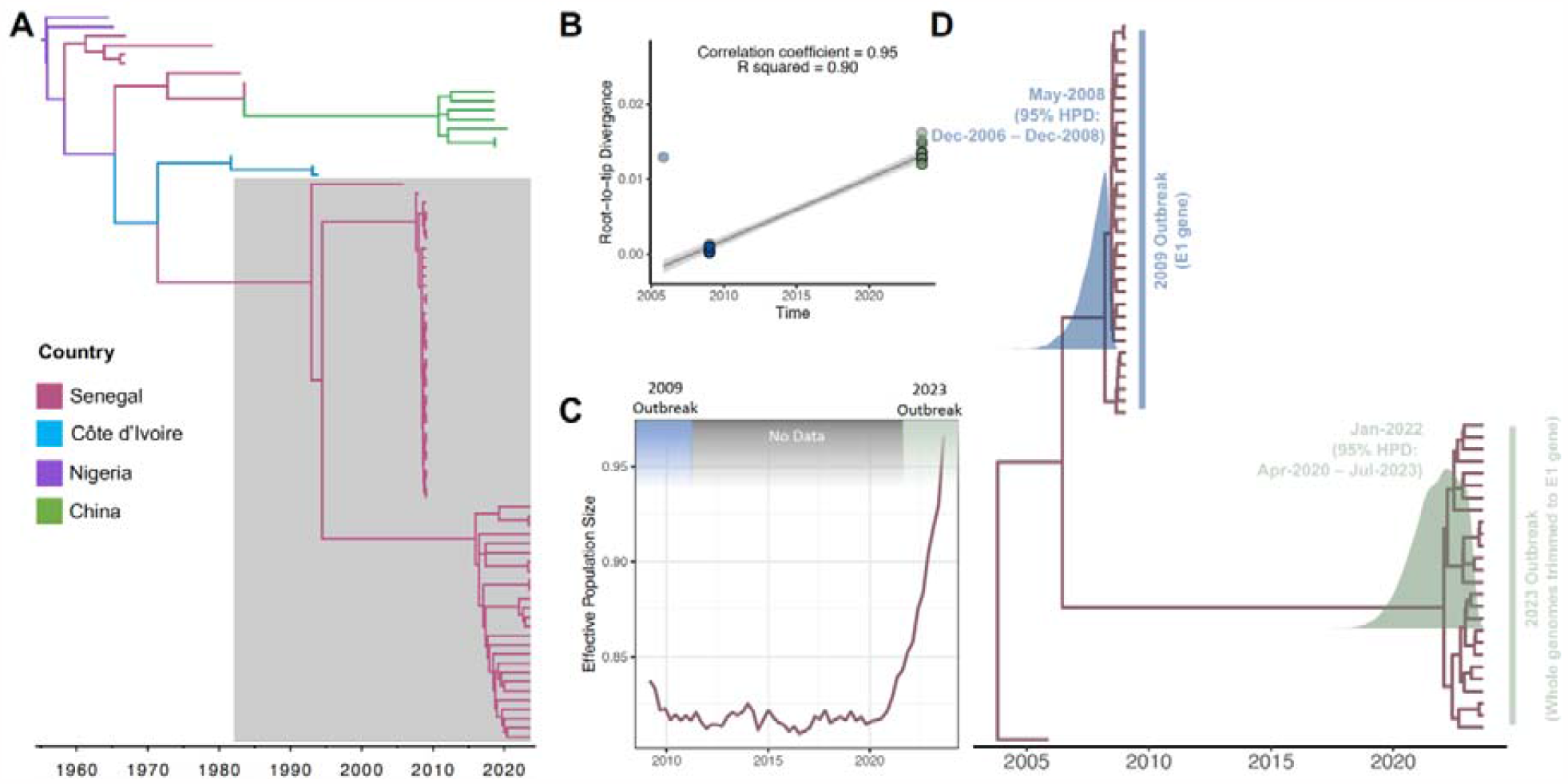
A) Timed maximum likelihood phylogeny of all West African genotype sequences of CHIKV. B) Root-to-tip repression containing sequences from the clade highlighted in gray in the previous panel, containing public CHIKV sequences from 2005, 2009, and newly sequenced ones from 2023. C) Effective population density estimates for CHIKV in Senegal. D) Maximum Clade Credibility (MCC) phylogenetic tree of 2005, 2009 and 2023 sequences, trimmed to E1 gene, of CHIKV in Senegal, showing the posterior distributions for the root node age of the 2009 and 2023 clades, with specified 95% HPD.

We worked closely with the district of Kédougou to investigate suspected cases in Ndiormi and Bandafassi (Figure 1C), cities close to Kédougou for CHIKV screening and characterisation of circulating lineages during this outbreak. The investigation started early in the outbreak in July with active case-finding, recording of symptoms such as fever, asthenia, myalgia, headache, arthralgia, and signs of bleeding over the past 30 days. Blood samples were collected from suspected cases and submitted to molecular screening. We obtained 57 plasma specimens from suspected cases in the Kédougou region (Figure 1A, C). The demographics of the sampled cases consisted of a majority of women over 30 years, and children and young adults of both sexes (Figure 1B, Supplementary Table S1). This seems to suggest infections were also acquired in households, therefore urban transmission, rather than solely sylvatic infections acquired close to forested environments. In contrast, investigations during the 2009 outbreak found no infection in <4 year olds, which seemed to indicate no household transmission^11^.

Kédougou region is a rural community located in South-Eastern Senegal close to the Niokolo-Koba National Park, with a population 133,487 people, 55% of whom are under 20 years old. The climate is Sudano-Guinean with a single rainy season (May-November). Widespread activities of hunting, logging and gold extraction are a source of human contact with the forest, which have typically been associated with sylvatic transmission of CHIKV^12^. Mean temperatures in Senegal have been steadily increasing, with 2023 set to be one of the hottest years on record (Figure 1D). By investigating the spatial trends of this temperature increase, we find that the Kédougou region shows the biggest rise in recorded temperature between the 1980s and the last decade (Figure 1E). Together with considerable forest cover in the region (Figure 1F), this could be enhancing the ecological suitability of large, sustained outbreaks originating from sylvatic transmission. The presence of vectors and a significant proportion of a young population in regular contact with non-human primates could further facilitate transmission of CHIKV from rural to urban areas in Senegal.

From the 57 plasma specimens collected, a total of 31 CHIKV positive samples were detected using RT-qPCR (S*eegene, Novaplex*^*TM*^ *Tropical fever virus Assay*), with CT values <45 (mean CT value = 27.15; range = 19.20, 40.90). In one patient, co-infection of CHIKV and DENV was detected via RT-qPCR, with CT values of 25.33 for CHIKV and 38.38 for DENV, respectively. Next-generation sequencing (NGS) was successful for all 31 samples, producing 15 near whole-genomes (>70% coverage) of CHIKV. In general, the lower the CT values (high viral loads), the more complete the genomic sequence (Supplementary Figure S1). All CHIKV genomes belonged to the West-African genotype, a lineage of CHIKV that has sporadically circulated solely in West Africa. These are the first locally-generated CHIKV whole genomes produced and publicly shared in real-time in Senegal and West Africa (GISAID EpiArbo^13^: EPI_ISL_18498014-EPI_ISL_18498039). The sequences are rather well conserved in the E1 gene, showing no known adaptation for vector competence^14^ (Supplementary Table S2), with the caveat that recent large CHIKV epidemics in Latin America also lack mutations thought to enhance vector competence^5^. CHIKV in Senegal has previously been isolated from *Aedes fucifer* mosquito species, known to be present in forest regions and to act as a bridge vector between sylvatic and urban cycles. No information is currently known about mosquito vectors involved in the 2023 transmission outbreak. The environmental factors that determine the risk of human infection, or the role of specific vector species in the transmission of CHIKV from sylvatic reservoir hosts to humans, remains unclear. Detection of the same CHIKV strain in *Ae. fuicifer* mosquitoes and non-human primate reservoirs would point towards this transmission being a result of a large sylvatic cycle. The detection of the virus in other *Aedes* species such as *Ae. Aegypti*, also present in Senegal^15^, would point to an urban transmission cycle.

Timed maximum likelihood phylogeny of all available CHIKV West-African genotype sequences (E1 gene or whole genomes) reveals that the 2023 outbreak sequences from Kédougou form a monophyletic cluster which is distinct from any previously identified strains in Senegal, and from other known strains of the West African genotype. The West African phylogeny is deep-rooted in West Africa, with approximate inferred root in the 1960s, coinciding with the first recorded spillover from enzootic cycles^2^. Recently isolated detection in China has been described before^16^. In order to understand viral emergence and transmission dynamics of this novel clade in Senegal, we employed a Bayesian phylogenetic reconstruction approach. Using a relaxed molecular clock model for the construction of a time-calibrated MCC phylogenetic tree, we infer the estimated time to most recent common ancestor (TMRCA) of the novel clade to be early 2022 (95% highest posterior density interval; HPD = April 2020 - July 2023), followed by a steady growth in effective population size until 2023 (the most recent sampling date in this study). The transmission clade associated with the 2009 outbreak has an estimated TMRCA dating back to early 2008 (HPD = December 2006 - December 2008).

This analysis supports the finding that the emerging clade circulating during the 2023 outbreak in Kédougou is phylogenetically distinct from the one characterized during the 2009 outbreak, suggesting a distinct spillover event from sylvatic cycles in Kédougou. The detection of sporadic CHIKV cases in the region in 2021 and 2022 supports the inference of the date of emergence of the circulating clade. However, it is not possible to confirm that these sporadic cases are basal to the 2023 outbreak clade without genomic data from this period. The lack of genomic data from the 2015 outbreak also makes it impossible to infer population dynamics between the two outbreaks. The last common ancestor of the two outbreak clades along with a CHIKV isolate sampled in 2005, existed before 2005 in Senegal and likely circulated in sylvatic cycles. This provides crucial insights into the progression of Chikungunya from emergence to escalating outbreaks. In both outbreaks (2009 and 2023), it appears that the virus emerged and circulated for ∼1 year before being detected in large outbreaks. The rural ecological setting of Kédougou, including close interaction between inhabitants and non-human primates, could facilitate spillover from sylvatic to urban cycles.

## Conclusion

This study highlights the emergence of a new clade of CHIKV within the West African genotype, discovered in real-time in Senegal during the current outbreak starting in July 2023. This clade was unrelated to strains sequenced in 2009, suggesting a likely novel spillover from sylvatic cycles. It appears that this new lineage circulated for at least one year before the current epidemic. Sequencing of additional CHIKV samples from 2021/2022 sporadic cases, and from other regions would be crucial to strengthen evidence of its origin and transmission dynamics. This need is underscored by the rising temperatures in Senegal, particularly in Kédougou, and the potential of seeding larger epidemics in urban settings, specially in light of the presence of low herd-immunity. The unprecedented experience of this real-time characterisation during the CHIKV epidemic in Senegal provide important insights to help public health response and future epidemic preparedness.

## Methods

### Study design and sampling

The investigation was conducted in collaboration with the Kédougou Health Centre in the Kédougou region (Ndiormi and Bandafassi) from 19 August to 23 August 2023. The following definitions were used during epidemic monitoring procedures. *Suspected case*: all persons residing in the surveyed areas of the Kédougou region who presented with fever (corrected axillary temperature ≥ 38°C), and at least two of the following signs: headache, myalgias, arthralgias, skin rash, retro-orbital pain; or haemorrhagic manifestations (epistaxis, gingivorrhagia, metrorrhagia, etc.) or neurological signs; *Probable case*: Any suspected case, living or dead, with an epidemiological link to a confirmed case of Chikungunya. *Confirmed case*: Any suspected or probable case of Chikungunya confirmed in the laboratory by ELISA test, IgM, RT-PCR, antigen detection, serum neutralization or viral isolation. In this cross-sectional study, 57 patients presenting with symptoms compatible with an Arboviral infection from the case definition were enrolled from the District of Kédougou in Senegal. The participants included 37 samples from Ndiormi and 20 from Bandafassi during the current outbreak. Blood samples were collected by the district’s staff. These samples were then sent to IRESSEF Diamniadio for confirmation and genomic sequencing.

### Ethical statement

This project was reviewed and approved by the Comité National d’Ethique pour la Recherche au Sénégal (CNERS) (Ref. N° 000031/MSAS/CNERS/SP/06/02/2023). All patients who provided consent to give samples for this study were enrolled and samples were collected by staff in the Kédougou district during the 2023 Chikungunya outbreak. The Direction de la Prévention, which is part of the Senegal Ministry of Health, was informed about IRESSEF intervention during Chikungunya outbreak in Senegal on August 8, 2023 with the reference number N/Réf: SM/IRESSEF/RARS/116.09.2023.

### RNA extraction and molecular screening

The samples were extracted using the MagMAX kit and processed on the Kingfisher platform according to the manufacturer’s instructions, with the samples being eluted in a volume of 50 μL. RT-PCR amplification was performed using the Novaplex™ Tropical Fever Virus Assay multiplex kit, which detects Chikungunya (CHIKV), Dengue (DENV), Zika (ZIKV), and West Nile (WNV) viruses (*Seegene, Novaplex*^*TM*^ *Tropical fever virus Assay*). Amplification was carried out on Bio-Rad’s CFX96 instrument, and the results were exported to the Seegene Viewer software.

### Setting up Whole Genome Sequencing at IRESSEF, Senegal

The CLIMADE consortium supports countries in Africa, Asia, and Latin America to rapidly respond to emerging vector-borne or waterborne disease outbreaks. The support provided includes training, the sharing of sequencing protocols, and when resources allow, the provision of reagents; and personnel to assist countries in rapidly characterizing and understanding emerging epidemics. Ongoing mentorship and assistance with data analysis are also provided as part of a weekly meeting where data analysis protocols are shared and discussed. This facilitates near real-time response to outbreaks to inform public health responses. In 2022, Dr. Padane, the first author of this paper, from IRESSEF, was awarded a long-term fellowship that enabled him to spend several months at the Centre for Epidemic Response and Innovation (CERI) at Stellenbosch University in South Africa to gain experience in whole genome sequencing (WGS) and epidemic response as part of CLIMADE consortium. Upon returning to Senegal, Dr. Padane initiated an integrated approach to enhance genomic surveillance and characterize the arboviruses in West Africa. The CERI team, the Fundação Oswaldo Cruz (FioCruz) team, and the KwaZulu-Natal Research Innovation and Sequencing Platform (KRISP) of the CLIMADE consortium supported Dr. Padane to to establish DENV and CHIKV surveillance in West Africa by providing training, protocols, reagents and consumables.

In short, the CLIMADE support process involved an assessment of IRESSEF laboratory capacity, which sequenced the samples locally in Senegal. The IRESSEF genomics laboratory contained all of the equipment for WGS sequencing using both Illumina and Oxford Nanopore Technologies sequencing. As the first cases of CHIKV were detected in Senegal on 17th July, Dr. Padane contacted the CLIMADE consortium for support on 17th August 2023, and the CERI and KRISP teams shipped the Illumina COVIDseq Test Assay and the Illumina Miseq v3 600 cycle Reagent kit to Senegal on the 15 September 2023. In addition, the shipment also included the CHIKV primers for amplicon sequencing, which a local company in South Africa synthesized. The shipment was received at IRESSEF on the 18th September 2023. The primer scheme for CHIKV sequencing, and other arboviruses, is available on the CLIMADE GitHub protocols (https://github.com/CERI-KRISP/CLIMADE/tree/master/Protocols/Arboviruses). The COVIDSeq protocol was adapted to perform the library preparation, followed by sequencing on the Illumina Miseq platform. This protocol is available on the CLIMADE GitHub website, and it is described in detail by *Elaine Vieira Santos, Debora Glenda Lima de La Roque in protocols.io (https://protocols.io/view/pathogen-whole-genome-sequencing-multiplexed-ampli-cgwbtxan)*. This protocol was modified to replace the DENV primers with those specific for CHIKV virus. The CLIMADE consortium provided advice and guidance during the library preparation and sequencing. The IRESSEF team was given guidance on the interpretation of the quality control and the calculations for the dilution of the final pooled libraries.

The resulting genomic sequences have been made available on GISAID Epi-Arbo database^13^ (EPI_ISL_18498014-EPI_ISL_18498039).

### Chikungunya alignment and Phylogenetic Analysis

CHIKV reference dataset was retrieved from two primary databases: the National Centre for Biotechnology Information (NCBI) GenBank and the Bacterial and Viral Bioinformatics Resource Center (BV-BRC) web interface (https://www.viprbrc.org). Any duplicate entries were removed. Sequence genotyping was performed using the Genome Detective Chikungunya Typing Tool^17,18^ (https://www.genomedetective.com/app/typingtool/chikungunya/). Sequences were then filtered for the West African Genotype (n = 115). The retrieved sequences were subjected to initial quality control to remove any unverified sequences and incomplete records (i.e geographical location and sampling dates; n=59). To ensure data uniformity and comparability, both newly generated CHIKV sequences (n = 31) and retrieved sequences (n = 55) were aligned against the CHIKV reference genome (NC_004162.2) using Nextalign v1.3.0 alignment tool^19^.

Maximum likelihood trees were generated using IQ-TREE v 2.2.2.2^20^ with 1000 bootstraps. The nucleotide substitution model TIM+F+I+R2 was selected according to Bayesian Information Criterion using Model Finder^21^. Molecular clock signal was evaluated using TempEst v1.5.3^22^. Potential outliers that violated the molecular clock assumption were removed prior to inferring time-scaled tree (TreeTime v 0.10.0^23^) using a molecular clock rate of 2.845 × 10^−4^ nucleotide substitutions per site per year.

### Phylodynamics Analysis

Time-calibrated Bayesian phylogenetic trees were constructed to estimate the time of emergence (time to most common recent ancestor, TMRCA) of the transmission lineage circulating during the 2023 outbreak, and the associated effective population size. The latest release of BEAST (v.1.10.4)^24,25^ was used along with the BEAGLE library v3.2.0^26^ to improve computational speed. The dataset for the analyses described here contain sequences from the 2023 outbreak from this study (n = 26 after discarding outliers) and publicly available Senegal sequences collected in 2005 (n = 1)^27^ and 2009 (n = 33)^11^ to root the tree. After alignment of this dataset (as described above), the sequences were trimmed to only retain nucleotide positions 10001 to 11293, corresponding to the E1 gene to be able to analyse what were originally whole genomes with E1 sequences from 2009. A relaxed clock model was applied for all analyses along with the HKY+G+I nucleotide substitution model. First, a constant population coalescent model assumption was used to estimate the TMRCA of two taxa groups: 2023 outbreak and 2009 outbreak. Secondly, a skygrid^28^ coalescent growth model with 64 grid points over the last 16 years (corresponding to grid points every 4 months) was used for effective population size calculations. In this second analysis, a normal prior was applied to the TMRCA parameter of each taxa group, centered on the TMRCA estimated in the first analysis with a standard deviation of 1. The purpose was to obtain a more precise estimation of the TMRCA of each taxa group while simultaneously reconstructing the effective population size. All BEAST runs were set up in BEAUti^29^, a user-friendly program for building phylodynamic models and specifying MCMC settings for a BEAST analysis. Subsequently, all analyses were performed in two independent runs of ∼500 □ million iterations each, or enough iterations to reach convergence. Convergence of MCMC chains was checked using Tracer v.1.7^30^ and ∼10% of initial chains was discarded as burn-in, or an appropriate number determined after visual inspection in Tracer^31^. Post-burnin samples were pooled to summarize parameter estimates using LogCombiner and TreeAnnotator^24^, including posterior probability of each parameter and maximum clade credibility (MCC) trees.

### Mutational Analysis

We evaluated mutation frequencies in different CHIKV genotypes by utilising CHIKV-GLUE (https://github.com/giffordlabcvr/CHIKV-GLUE), a sequence-oriented resource for CHIKV developed using the GLUE software framework^32^. CHIKV-GLUE comprises a relational database capturing all CHIKV sequences accessible in GenBank up to November 1, 2023. Genotypes were assigning to CHIKV sequences using GLUE’s maximum likelihood clade assignment (MLCA) algorithm^32^. Isolate-specific information, including country and year of sampling, as well as the isolation host species, were extracted from GenBank XML files via GLUE’s ‘GenBankPopulator’ module. All sequences contained within CHIKV-GLUE are aligned to one another, with alignment information being stored in the relational database. Multiple sequence alignments (MSAs) were created using a codon-aware alignment algorithm integrated into the GLUE framework and based on the Basic Local Alignment Search Tool (BLAST)^33^. For each CHIKV genotype, we constructed a dedicated MSA wherein the coordinate space is restricted to a genotype-specific reference sequence, annotated with the coordinates of individual CHIKV genome features. The accession numbers for constraining reference sequences are as follows: Asian, HM045813; ECSA, FJ445426; West African, HM045785. We calculated mutation frequencies in each genotype-specific MSA by using GLUE’s command layer to interrogate the CHIKV-GLUE database. The ‘amino acid frequency’ command provided the frequency of all amino acids in a specified MSA, with coordinates referenced to specific genome features in the constraining reference sequence.

## Data Availability

The resulting genomic sequences have been made available on GISAID Epi-Arbo database (EPI_ISL_18498014-EPI_ISL_18498039).

## Acknowledgement

This work has been supported by CLIMADE (Climate Amplified Diseases and Epidemics), the National Institute of Health USA (U01 AI151698) for the United World Antivirus Research Network (UWARN), Abbott Laboratories and by NIH NIAID U01AI151698, United World Antiviral Research Network, part of the NIAID CRIED network and WANETAM (West African Network in Tuberculosis AIDS and Malaria). Data analysis support was provided by the Centre for Epidemic Response and Innovation (CERI) at Stellenbosch University. Reagents for WGS were provided by CERI’s CLIMADE grant from the Rockefeller Foundation (HTH 017). The authors would like to express their gratitude to the Ministry of Health and to Drs. Mamadou Ndiaye, Boly Diop for their support. The authors would like to express their gratitude to the Kédougou District team particularly Dr Fodé Danfakha for significant contribution and assistance during this project. The authors would like to express their recognition to the IRESSEF team and CERI group for their commitment during this project and the patients for their participation in this work.

