## Supplementary Material for "An emerging clade of Chikungunya West African genotype discovered in real-time during 2023 outbreak in Senegal"

**Supplementary Materials**

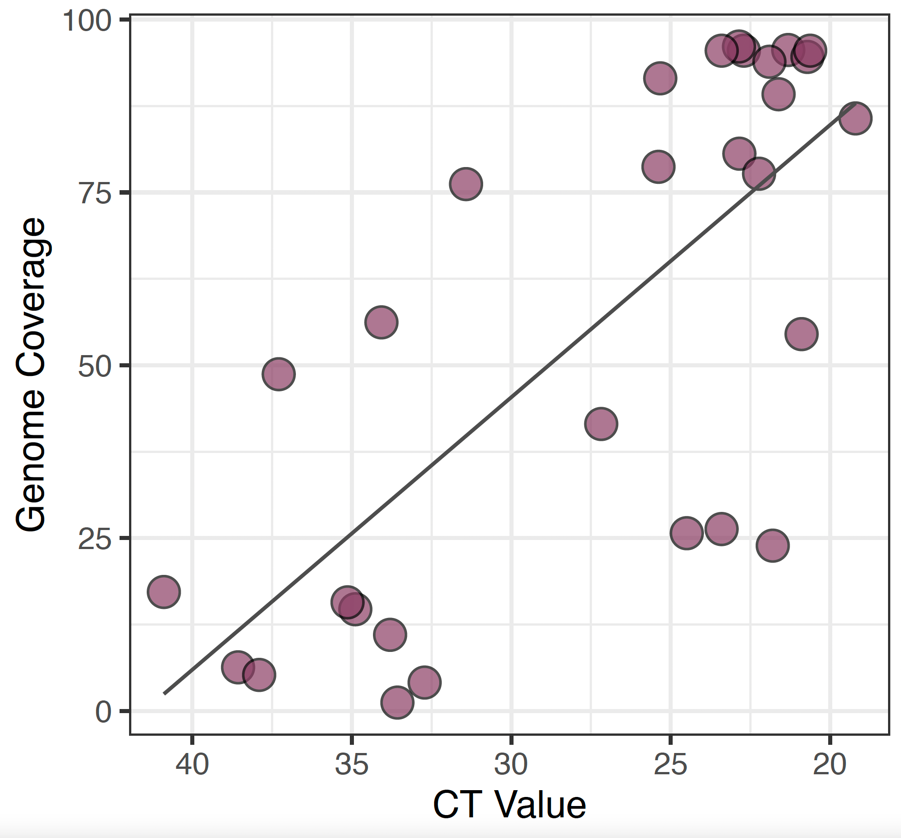

**Supplementery Figure S1.** Sequencing output from specimens testing positive with various CT values, indicating high genome coverages obtained with lower CT values (higher viral loads)

**Table S1**. Demographic characteristics of study population.

| Sampling locations | Bandafassi | | | Ndiormi | | |
| --- | --- | --- | --- | --- | --- | --- |
| Features | CHIK positive | CHIK negative | P | CHIK positive | CHIK negative | P |
| Serological_Tests (IgM) | (n=2) | (n=10) |  | (n=15) | (n=6) |  |
| RT-PCR_Chikunguya (Tropical Fever Kit) | (n=10) | (n=8) |  | (n=21) | (n=16) |  |
| Age in year, median | 8 | 17 | 0,37 | 15 | 20 | 0,28 |
| Gender (% male) | 60% | 25% | 0,31 | 81% | 56% | 0,2 |

**Table S2: Frequencies of mutations associated with vector adaptation in the E1 gene**

| **Feature** | **Codon** | **AminoAcid** | **Asian Lineage** | | **ECSA and ECSA-IOL** | | **West African** | |
| --- | --- | --- | --- | --- | --- | --- | --- | --- |
|  |  |  | **No of Sequences** | **Frequencies** | **No of Sequences** | **Frequencies** | **No of Sequences** | **Frequencies** |
| E1 | 226 | A | 803 | 99.75 | 3050 | 74.19 | 144 | 100.00 |
| E1 | 226 | G | 2 | 0.24 | 0 | 0 | 0 | 0 |
| E1 | 226 | L | 0 | 0 | 1 | 0.02 | 0 | 0 |
| E1 | 226 | V | 0 | 0 | 1060 | 25.78 | 0 | 0 |
| E1 | 80 | V | 742 | 100.0 | 3321 | 99.31 | 142 | 100 |
| E1 | 80 | A | 0 | 0 | 12 | 0.35 | 0 | 0 |
| E1 | 80 | L | 0 | 0 | 9 | 0.26 | 0 | 0 |
| E1 | 80 | P | 0 | 0 | 2 | 0.05 | 0 | 0 |
| E1 | 82 | T | 742 | 100.0 | 3489 | 99.34 | 142 | 100 |
| E1 | 82 | A | 0 | 0 | 6 | 0.174 | 0 | 0 |
| E1 | 82 | I | 0 | 0 | 16 | 0.45 | 0 | 0 |
| E1 | 82 | S | 0 | 0 | 1 | 0.02 | 0 | 0 |
| E1 | 84 | V | 742 | 100.0 | 3590 | 99.58 | 142 | 100 |
| E1 | 84 | D | 0 | 0 | 13 | 0.36 | 0 | 0 |
| E1 | 84 | F | 0 | 0 | 1 | 0.03 | 0 | 0 |
| E1 | 84 | I | 0 | 0 | 1 | 0.03 | 0 | 0 |
| E1 | 98 | A | 0 | 0 | 3374 | 88.05 | 145 | 100.0 |
| E1 | 98 | T | 775 | 100.0 | 457 | 11.93 | 0 | 0 |
| E1 | 98 | V | 0 | 0 | 1 | 0.0265 | 0 | 0 |
| E1 | 211 | E | 805 | 99.87 | 1448 | 35.19 | 0 | 0 |
| E1 | 211 | K | 1 | 0.12 | 1498 | 36.41 | 145 | 100 |
| E1 | 211 | G | 0 | 0 | 1 | 0.02 | 0 | 0 |
| E1 | 211 | I | 0 | 0 | 10 | 0.24 | 0 | 0 |
| E1 | 211 | L | 0 | 0 | 1 | 0.02 | 0 | 0 |
| E1 | 211 | N | 0 | 0 | 69 | 1.67 | 0 | 0 |
| E1 | 211 | R | 0 | 0 | 2 | 0.04 | 0 | 0 |
| E1 | 211 | T | 0 | 0 | 1085 | 26.3 | 0 | 0 |
